# Suppressing the impact of the COVID-19 pandemic using controlled testing and isolation

**DOI:** 10.1101/2020.05.03.20089730

**Authors:** Kobi Cohen, Amir Leshem

## Abstract

The Corona virus disease 2019 (COVID-19) has significantly affected lives of people around the world. Today, isolation policy is enforced by identifying infected individuals based on symptoms when these appear or by testing people and quarantining those who have been in close contact with infected people. In addition, many countries have imposed complete or partial lock-downs to control the spread of the disease. While this has resulted in some some success in slowing down the spread of the virus, lock-downs as well as widespread quarantine have devastating effects on the economy and social life. Thus, governments are urgently looking for efficient strategies to significantly relax lock-downs, while still controlling the spread of the virus. We argue that this can be done by using active feedback to control testing for infection by actively testing individuals with a high probability of being infected. We develop an active testing strategy to achieve this goal, and demonstrate that it would have tremendous success in controlling the spread of the virus on one million people, using 3,000 tests per day. Our results show up to a 50% reduction in quarantine rate and morbidity rate in typical settings as compared to existing methods.

## Introduction

The outbreak of the COVID-19 has revealed the widespread effects a pandemic can have on all spheres of life from health, to social life, to the economy (31). The main thrust of efforts to control the spread is to decrease the reproduction rate to flatten the curve of the total number of infected individuals per day. This is absolutely necessary to reduce the load on the health system, although the infection may spread in the population over an extended period of time (1, 2).

The most widely implemented response on the part of the international community to the exponential growth of the infection has been widespread quarantine and lock-downs (32,37). While isolating people is an effective tool to decelerate the spread, imposing a complete quarantine for everyone for a relatively long period of time until the virus is suppressed is impractical primarily because of its devastating effects on the economy and social life. Moreover, if proper population monitoring is not enforced there may be further waves of the disease.

Since imposing significant (or even complete) quarantine can have devastating economic consequences, early detection of Corona positives is of paramount importance to suppress the spread of COVID-19 as early as possible. Hence, the second component in fighting the pandemic is to devise and manufacture COVID-19 tests. Today, health systems around the world prioritize the administration of tests to people who have had direct contact with an infected person or who are symptomatic. Some countries, such as South Korea, are implementing extensive resources to test people widely and randomly to detect areas where the pandemic is likely to spread. Nevertheless, these strategies do not exploit resources effectively. First, only testing symptomatic people results in a loss of precious time, during which the individual can infect others before symptoms appear. Furthermore, many people (about 10% – 30%) are asymptomatic, and thus cannot be identified using this strategy. In addition, testing the entire population on a daily basis to detect people who remain asymptomatic is impossible because the number of tests that labs can either produce or process is limited. Clearly, quarantining the entire population for a long period of time has devastating effects on economy and social life. **For these reasons, we address the imperative question of whether a strategy to test and isolate a relatively small number of people while effectively suppressing the spread of COVID-19 can be developed. Specifically, the main challenge is how to efficiently test and isolate people in the population to significantly reduce the negative impacts of the spread of the virus, in terms of the burden on the health system (i.e., flattening the curve of infected people per day), total morbidity (i.e., reducing the total number of severe COVID-19 patients), and the economic and social impact (i.e., reducing the total number of people required to stay in quarantine and the time people spend in quarantine). In this paper, we address this challenge, and develop active testing and isolation (ATI) strategy that achieves this goal**.

The mitigation of a pandemic can be achieved using a range of interventions to reduce transmission (16). Recently, partial lock-down strategies have been suggested, in which half of the population remains under lock-down, where the other half is released, in alternation (24), or by applying a cyclic schedule of 4 days of work and 10 days of lock-down (19). However, these strategies lead to more than a 50% loss in economic productivity, which we aim to avoid. These strategies are based on the effectiveness of quarantine, and are simple to analyze. However, they do not consider wide-scale testing, which is available. By tracing and quarantining only those at risk (e.g., those in close contact with an infected individual, depending on the duration of contact and distance), epidemics can be contained without using aggressive lock-downs (7,11), using surveillance and containment measures for patients (26), contact tracing (8,10) and mathematical modelling of the transmission dynamics of the infection (22).

As reported recently in (29), increasing the COVID-19 test rate per population would have a significant positive impact on mitigating the pandemic around the world. The missing component in most existing methods is that they do not use COVID-19 tests to identify infected people in development and analysis of strategies to suppress the pandemic. However, there are a number of recent studies that incorporate tests to mitigating the spread (29), and provide worst-case analysis of a risk-based selective quarantine (33). Nevertheless, these methods use passive testing in the sense that post-processing the test results is not used to improve the future sampling process of the tested population (i.e., open-loop). In a recently published *Science* paper (7), Ferretti et al. concluded that viral spread could be controlled using contact-tracing by testing close contacts of positive cases. In this paper, we argue that suppressing viral spread can be accomplished much more efficiently by developing a controlled sensing methodology for contact-tracing, rather than testing close contacts in a deterministic manner as in (7). *Controlled sensing*, a.k.a active sensing, is based on classic sequential experimental design theory (3,30), and has attracted growing attention in recent years in various hypothesis testing and dynamic search problems (4, 14, 17, 18, 20, 25, 27, 34, 38). Controlled sensing policies, where the location of the sensing is judiciously selected, have also been used to identify influence in social networks (36), as well as to learn the dynamics in general networks such as gene networks and social networks to reduce the overall number of required experiments (35). **The basic idea behind controlled sensing theory when translated to the realm of detecting COVID-19 patients is that the learner (i.e., the testing system) can use the results obtained from previous tests to adjust its monitoring strategy in a closed-loop manner and improve its future testing and learning process. Intuitively, as more tests are performed, the learner becomes more certain about the true state of the population, which in turn leads to better choices of people to test**. This approach was shown to achieve the asymptotic information theoretic bound in various hypothesis testing and dynamic search problems as the error probability approaches zero (see e.g., (15, 25, 27, 28) and our previous work (4, 5, 12, 17)). Here, we develop a controlled testing methodology to control the spread of the COVID-19 pandemic. We report a significant reduction (up to 50%) in both quarantine rate as well as morbidity rate in typical settings of COVID-19 parameters as compared to existing studies.

## Main results

In this paper we develop a controlled testing scheme for better containment of the spread of the COVID-19 pandemic. Controlled testing is based on using the results of prior tests to decide on the proper test to be conducted next. This method can significantly enhance the control of the spread of pandemics as well as future epidemics. Furthermore, its association with accurate technological contact tracing techniques can prove extremely efficient in preventing the recurrence of pandemics when international travel will start up again, since it can significantly accelerate the identification and prevention of the spread via new cases.

The technique exploits a large scale agent-based stochastic model, where we monitor our belief regarding each agent’s state and select to test for infection the most probable agents to be infected based on belief and the total number of tests available. Based on this model we optimize controlled tests based on belief estimated from contact tracing data as well as symptom reporting. We demonstrate the effectiveness of the technique using a stochastic spread model in populations of 100,000 and 1 million people. We show that we can reduce the quarantine period (defined as the period where more than 1% of the population is quarantined) by up to 50%, while reducing the peak of number of infected people per day, as well as the total morbidity by a significant amount. Our methodology allows optimal exploitation of the testing capacity, and the method can be implemented under testing capacity limitation. With a capacity of 0.3% of the population of an infected region, we already show significant benefits.

The importance of these numbers is not only in coping with the first wave of the pandemic, but also monitoring new occurrences of the disease over time since it will significantly reduce the overall quarantine rate and accelerate the containment time in these future waves.

This large scale model differs substantially from mean-field differential equations based epidemiological models in that it allows to microscopically track the target individuals, something which cannot be done using generalizations of SIR, SIRS or other common epidemic spread models which are based on large population mean-field techniques (6, 9, 13, 21). It can also easily incorporate new models for infection probability. For example, a better understanding of the spread when people use masks or use other protective methods can be quantified and used to update the probabilities accordingly. Similarly, the model can be used to more accurately exploit the duration of contact provided by contact tracing measures. This can be easily incorporated as another factor into the model.

### Performance evaluation

We begin by describing simulated results that support our claims. Then we provide the details of the proposed active testing and isolation (ATI) (control algorithm) and end with the specifics of the stochastic agent-based model and the related algorithms used to direct the active testing process. We present three sets of results. In the first set we study the possibility of quickly controlling an outbreak in its early phase. Then, in the next two sets, we discuss the amount of testing required to suppress an advanced outbreak with 0.1% of the population already infected.

As a baseline to our study we consider three methods: (a) Isolation without testing (NTI) where only symptomatic people are tested. This was the approach, e.g., in Israel; (b) widespread random testing and isolation (RTI), i.e, following the NTI policy and augmenting it with widespread tests for anyone interested in getting tested; and (c) deterministic testing and isolation (DTI) based on contact tracing as presented in Ferreti et al. (7). In this technique, when an infected agent is detected, all its close contacts are quarantined and are told to get tested. While DTI is much more efficient than the previous two methods, it allocates efforts to people unlikely to be infected, either because of the duration of contact or other prior information collected by the contact tracing. It also incurs a delay in testing second order infections. We propose two levels of active testing: Computing infection probabilities based solely on first order contacts or by solely using second order contacts, dubbed 1st-order, and 2nd-order ATI, respectively. The latter becomes more efficient when the testing capacity is large.

#### New eruption in a small city

To simulate the case of an early outbreak, we consider the case of 50 infected people in a city of 100,000 people. We also use this case to test the magnitude of success of the quarantine. We assumed that false negative of tests are 10% and changed the quarantine success rate from 0.7 to 1. We assumed that we have 300 tests available for testing the population in this city every day, which is 0.3% of the population. This is a small number which can be easily conducted in cities of this size. First we present the performance of the various techniques with respect to a quarantine success rate of 70% in Fig. 1.

**Figure 1:**
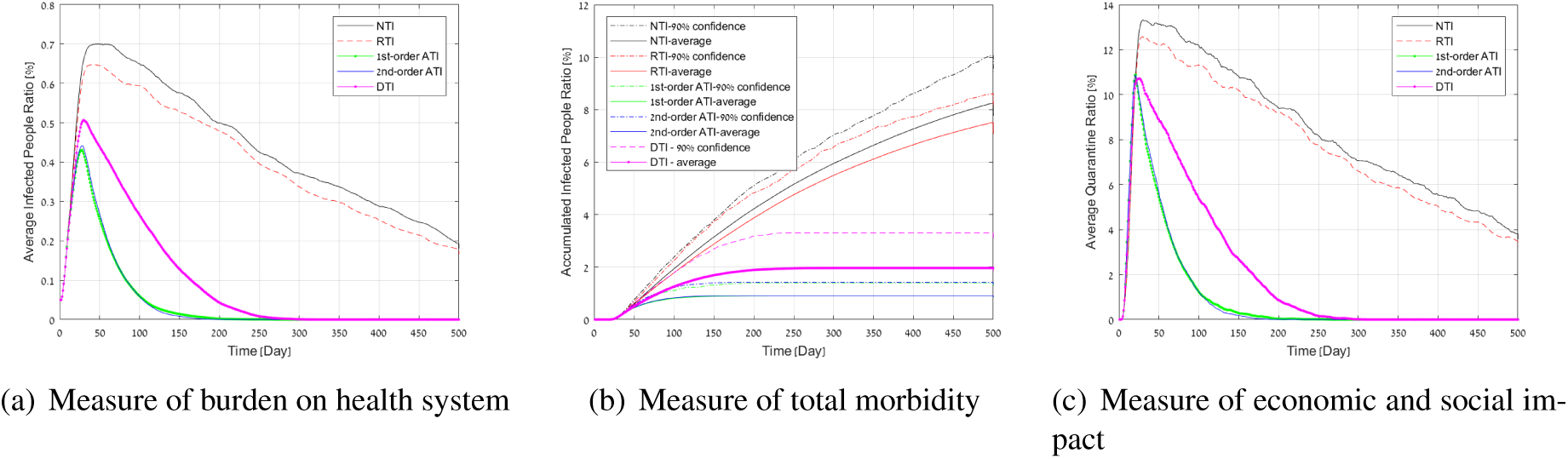
Simulation results for a COVID-19 outbreak in a population of 100,000 people, and a low testing capacity (0.3% per day).

As shown, as long as the tests are properly controlled by the ATI, the decay time is relatively short with a significantly lower quarantine rate even when the quarantine is only observed by 70% percent of the population required to be in quarantine. In contrast, with such a low number of tests, the RTI is practically equivalent to the NTI. The combination of a low quarantine rate and random testing is incapable of providing an effective means to block the spread, although it does manage to flatten the curve over a very long period. DTI has better performance, and the active approach is significantly superior in controlling future small outbreaks (DTI requires twice quarantine days, and results in twice morbidity rate, and higher load on health system as compared to ATI). Thus, accurate contact tracing with controlled testing does not require a huge amount of testing to stop the outbreak. **We observed (see supplementary material for detailed numbers) that NTI achieves results that are between tens and up to hundreds of percent worse than ATI when the quarantine success rate is between** 100% **and** 70%**, respectively**. Tables 1, 2 present the total infected population, overall quarantine days over the epidemic and the peak infection for all methods for quarantine success rates of 70%, 80%, respectively. It can be seen that the total days lost due to quarantine by NTI are 4 times and up to 8 times greater than ATI, and even DTI requires 40% and up to 100% more quarantine days than ATI, mainly because it does not order people to be tested according to the probability of infection. NTI and RTI also result in 4 times and up to 9 times more infected individuals.

**Table 1:** Performance evaluation for a 70% quarantine success rate

| Measure/method | NTI | RTI | DTI | 1st-order ATI | 2nd-order ATI |
| --- | --- | --- | --- | --- | --- |
| Total # of infected people | 8,256 | 7,516 | 1,962 | 909 | 911 |
| Peak # of infected people | 701 | 648 | 507 | 431 | 442 |
| Total # of days in quarantine | 4,118,872 | 3,816,708 | 1,046,777 | 509,401 | 508,265 |

**Table 2:** Performance evaluation for an 80% quarantine success rate

| Measure/method | NTI | RTI | DTI | 1st-order ATI | 2nd-order ATI |
| --- | --- | --- | --- | --- | --- |
| Total # of infected people | 2,042 | 1,658 | 620 | 445 | 432 |
| Peak # of infected people | 509 | 488 | 408 | 349 | 344 |
| Total # of days in quarantine | 1,298,703 | 1,055,431 | 395,188 | 286,326 | 278,950 |

#### Developed outbreak with 0.3% testing capacity

We now test the results of controlling an outbreak with 0.1% of the population already infected for a region of one million people. First we assume a testing capacity of 3,000 tests per day, which corresponds to 3,000 daily tests. We assumed a false negative rate of 20%. The values of 10 – 20% are typical to current testing methodology, with some low quality testing kits presenting worse performance. In Fig. 2(a), we present the average number of infected people as a function of time (in days). In Fig. 2(c), we present the average number of people in quarantine as a function of time. The average accumulated number of infected people with time is presented in 2(b). In addition to the average performance, in Fig. 2(b) we present the upper confidence of 90% to demonstrate the significance of the results in terms of the morbidity in the population under the worst 90% realizations that we obtained. To simplify the readability of the results, we did not present this measure in Figs. 2(a), and 2(c). Nevertheless, we obtained similar significance in these measures as well.

**Figure 2:**
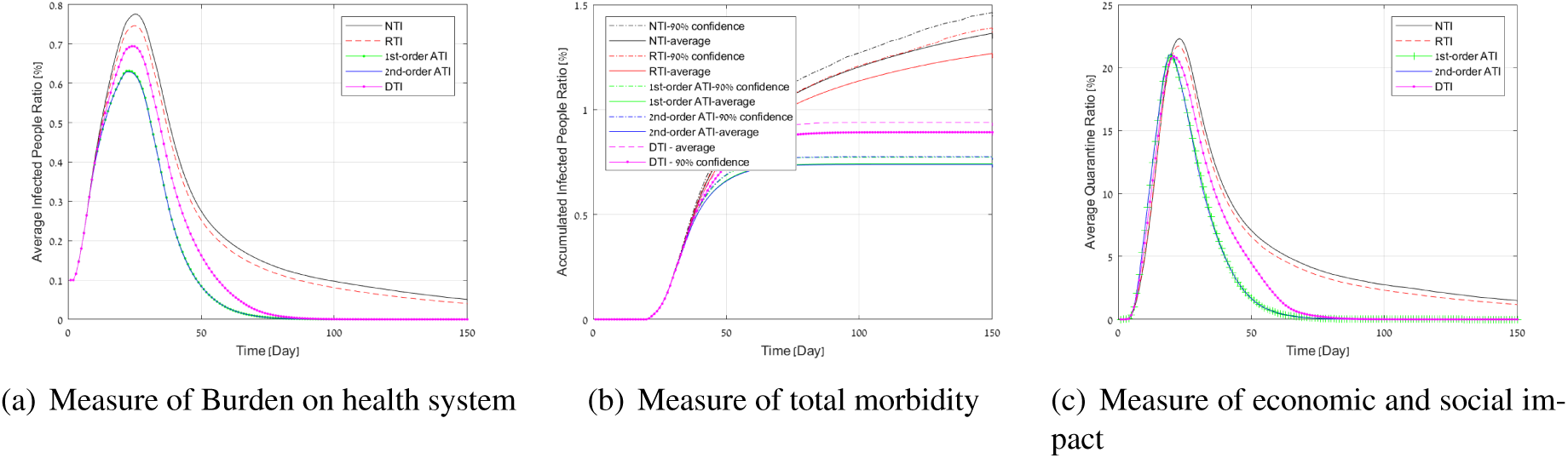
Simulation results for a COVID-19 outbreak in a population of 1 million people, and a low testing capacity (0.3% per day).

The figures show that NTI performs the worst on all performance measures, whereas RTI achieves slightly better performance than NTI, and DTI is better than both. All these methods are inferior to ATI. These results support the observation that a small number of random tests cannot achieve a significant improvement in controlling the spread of COVID-19, while the suggested ATI strategy (both first and second-order) succeeds in doing so. Specifically, the peak load in Fig. 2(a) under NTI and DTI, respectively, is 25%, and 11% higher than ATI. The total morbidity in Fig. 2(b) under NTI and DTI, respectively, is 85%, 20% higher on average, and 100%, 20% higher on 90% upper confidence than ATI. The quarantine curve in the 1% quarantine ratio in Fig. 2(c) under NTI and DTI, respectively, is more than three times, and 1.2 times wider than the quarantine curve under ATI. These results demonstrate the importance of testing people actively based on past observations in addition to the isolation strategy. It can be seen that first-order and second-order ATI achieves roughly the same performance in this case. This can be explained by the fact that when the number of tests is relatively small, both strategies give high weight to testing first-order neighbors. Thus, in this case, the first-order approximation is sufficient for effective active testing.

#### Developed outbreak with 5% testing capacity

We now study the effect of massive testing. To that end, we assumed up to 50,000 tests per day. We ran 100 Monte-Carlo trials, each with a random graph realization of one million people. Fig. 3(a) depicts the average number of infected people as a function of time (in days). The average accumulated number of infected people with time is presented in 3(b). Fig. 3(c), depicts the average number of people in quarantine as a function of time.

It can be seen that NTI performs the worst in all performance measures, whereas RTI achieves a significant improvement over the previous experiment. This is expected, since increasing the number of tests contributes to the detection of randomly selected infected people. DTI now coincides with first order active testing. This is mainly because with such a large number of tests, all first order neighbors can be tested. However, in this case, the second order active testing yielded significant gains. Specifically, the peak load in Fig. 3(a) under NTI and DTI, respectively, is 200%, and 18% higher than 2nd-order ATI. The total morbidity in Fig. 3(b) under NTI and DTI, respectively, is 225%, 20% higher on average, and 330%, 20% higher on 90% upper confidence than 2nd-order ATI. The quarantine curve in the 1% quarantine ratio in Fig. 3(c) under NTI and DTI, respectively, is more than three times, and 1.5 times wider than the quarantine curve under 2nd-order ATI. These results support the observation that increasing the number of tests increases the performance gain of the second-order ATI over the first-order ATI, since the second-order ATI seeks out infected people who are located outside the first-order neighborhood more efficiently.

**Figure 3:**
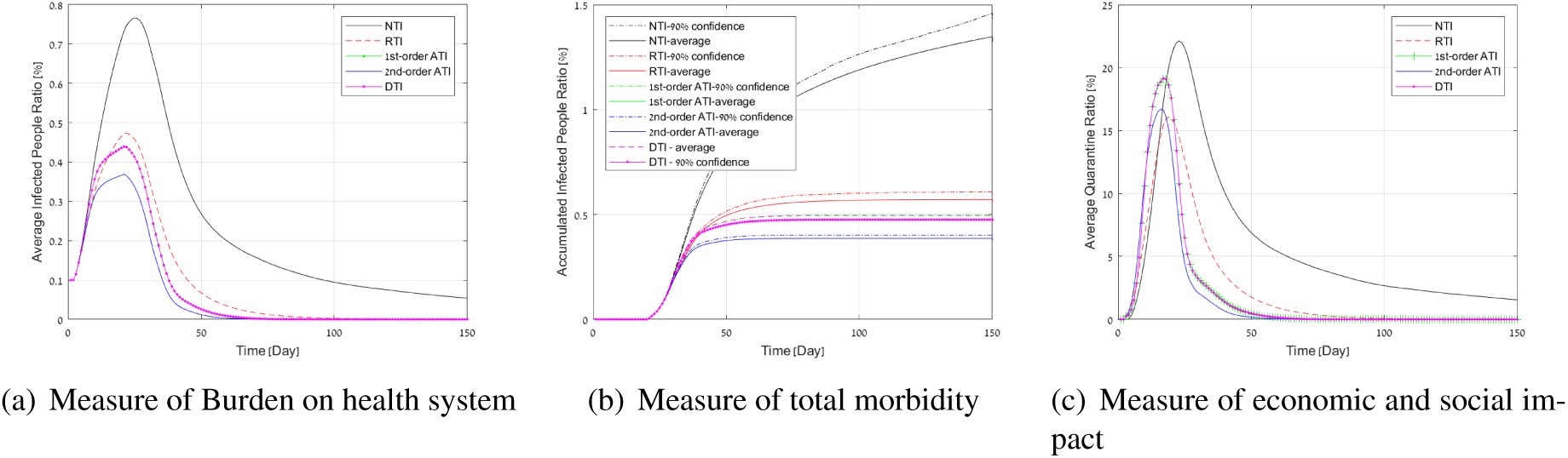
Simulation results for a COVID-19 outbreak in a population of 1 million people, and a high testing capacity (5% per day).

### Description of the active testing and isolation (ATI) algorithm

We now describe the proposed active testing and isolation (ATI) algorithm which controls the testing process. Let *N_T_* be the total number of test labs available for testing a population in a given region. Let *p_j_* be the estimate probability of agent *a_n_* being infected (i.e.*a_n_*, belief), and 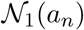 be the first-order neighbors (i.e., close contacts) of agent *a_n_* (the specifics of the model are given in the next section). Every day (or any other unit of time) the ATI algorithm works as follows:

1. **Identifying infected people and isolating them:** For the released population, check whether there are new infected people based on infection symptoms or preceding test results if tested. Send those infected people to quarantine until recovered. Denote these people by 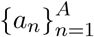.
2. **Isolating first-order neighbors in quarantine:** Order the first-order neighbors 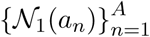 into quarantine for at least 14 days. Release them if symptoms have not appeared, or they have recovered, or if test results were negative if tested.
3. **Updating the belief over the population graph:** Update the beliefs p_j_ for all people in the population graph based on new information (e.g., identifying new infected people, isolating new people).
4. **Actively testing the most likely infected people:** Test all people with top *N_T_* beliefs among the entire population, regardless of their symptomatic state.
5. **Repeat:** Go back to Step 1 (for instance, a single day ended).

Note that the ATI algorithm tests and isolates people actively in a closed-loop manner. Steps 1, 2 identify infected people based on symptoms or test results. Then, people who are declared infected and their first-order neighbors are quarantined. The outcome of these steps is used to update the beliefs of all people in the population graph, which is then used to sample and test the most likely infected people regardless of their symptomatic state. This is the crucial concept that enables ATI to identify infected people early before symptoms appear, and decreases viral reproduction.

## Stochastic model for controlled testing

The proposed controlled testing scheme requires an underlying stochastic model to be able to use the current testing results to estimate the likelihood of a person being infected given the test results. We used a microscopic stochastic model which includes every agent in the population as described below. We model the connections between all *N* people in an area (e.g., state, district, city) as an undirected graph, where the people are represented by a set 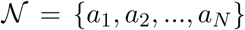of agents (or nodes or vertices), and ”close contacts” between people who might cause infection are represented by a set *E* of edges^1^. The existence of an edge (*n, i*) ∈ *E* between persons *n*, and *i* is determined based on criteria for potential infections. For example, being within 2 meters of a COVID-19 patient for more than 15 minutes is defined as close contact by the Israeli Ministry of Health, which can be represented by an edge in our population graph model. By using more accurate distance determination provided by short range wireless communication such as Bluetooth we might be able to define or learn a probability distribution from the data for the probability of infection. This turns the graph into a weighted graph, with time varying weights.

For each agent *a_n_* we define its set of first-order neighbors by:

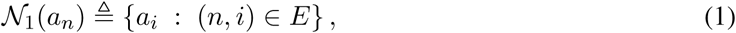

which represent all close contacts with respect to agent *a_n_*. Similarly, we define the second-order neighbors of an *a_n_* by:

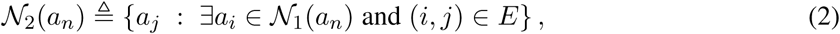

which represent all close contacts with respect to agent *a_n_*’s first-order neighbors. We focus only on first and second neighborhoods of agents. Nevertheless, higher neighborhood orders can be defined similarly. Intuitively, if agent *a_n_* is infected, then the probability of him or her infecting other people over the graph decreases as the neighborhood order increases.

We also allow for external independent information regarding each individual’s probability of being infected based on physical measurements, such as temperature or daily reporting of symptoms, which can be easily factored into the agent state.

Each agent has two local states; namely, a physical state *S_P_*(*a_n_*), and controlled state *s_C_*(*a_n_*). The physical state specifies its health condition (healthy *S_H_* /infected *S_I_* /recovered *S_R_* /dead *S_D_*), whereas the controlled state specifies whether the agent is in quarantine (so he or she cannot infect or be infected) or released. The controlled state takes one of two possible values: *s_Q_* (quarantine), which represents a person who was ordered to be in quarantine and *S_F_* which denotes a person out of quarantine.

The active testing algorithm has prior information regarding each agent’s state described by a probability distribution over the four possible values of the physical states, and knowledge of the control state.

The algorithm observes the physical state of the person, either by testing for COVID-19 or by the emergence of symptoms. This observation of *a_n_* is denoted by *o*(*a_n_*), which equals one if *a_n_* was confirmed infected. Otherwise, it equals zero. Note that observations might result in errors when inferring the physical states; for example, due to false negatives and false positives in COVID-19 tests. These considerations are taken into account in our simulation environment.

### Infections over the population graph

Contagions between people are probabilistic in the stochastic population graph. The probability that infected agent *a_n_* infects 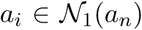 through path (*n, i*) ∈ *E* is denoted by *P*_(*n,i*)_. Typically, *P*_(*n,i*)_ is set such that the average number of individuals a patient infects is given by the reproduction rate.

The key in operating efficient population testing to stop the spread of the virus is to test the people most likely to be infected (so we can identify them before symptoms appear). The estimated probability of person *a_n_* being infected is denoted by *p_n_*, which we refer to as a *belief*. Let 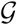 be the graph topology, including the infection probability rules over the graph. Let 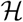 be the history of the revealed knowledge by the algorithm up to the current time, including all controlled states, and observations. Then, at each given time the ATI algorithm updates the beliefs based on 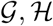:

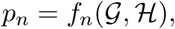

for all *n* = 1,…, *N*.

Note that the beliefs are updated temporally by taking into account practical aspects of COVID-19 effects on patients. For example, the probability of a second-order neighbor of an identified patient (who was quarantined when detected) being infected decreases with time as long as symptoms do not appear. Therefore, the beliefs must be decreased for all people who are not identified at each time step. Furthermore, the belief of a person that was tested and obtained negative test results should be decreased. This side information is captured by the history 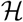. A detailed development of the belief update is given in the supplementary materials.

An illustration is provided in Fig. 4, where the population graph contains two infected patients *a_n_*, and 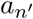. Given that these patients are identified, we can use the graph topology, and the infection probability over the edges to estimate the probability that *a_j_* is infected. Furthermore, assuming that 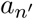 has an equal probability to infect his or her first-order neighbors *a_j_, a_m_, a_k_*, we can further infer that it is more likely that *a_j_* is infected than *a_m_* is (since person *a_j_* is a first-order neighbor of patient 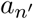, as well as a second-order neighbor of patient *a_n_*, whereas person *a_m_* is a first-order neighbor of patient 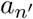, and only a fourth-order neighbor of patient *a_n_*), i.e., *p_j_* > *p_m_*. Therefore, applying efficient controlled testing strategy should prioritize testing *a_j_* over *a_m_*.

**Figure 4:**
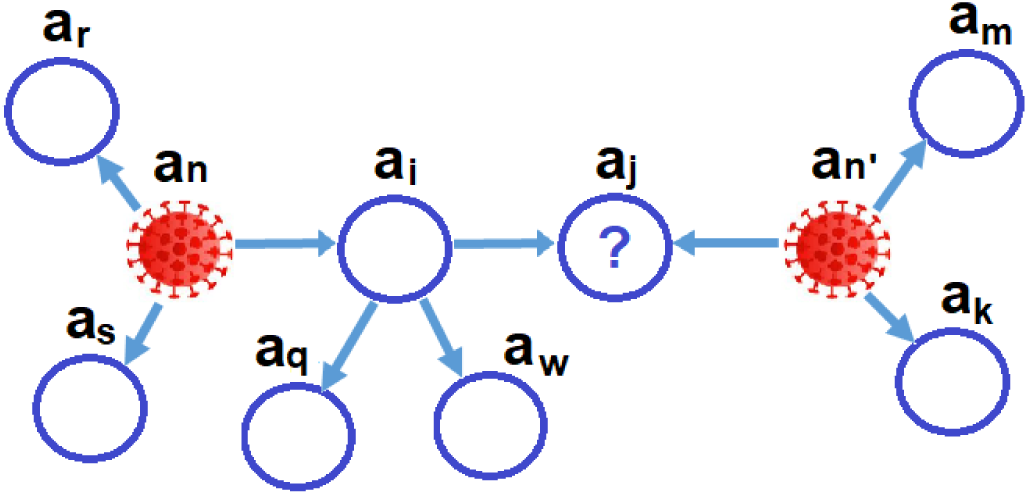
An illustration of infections over the population graph. Person *a_j_* is a first-order neighbor of patient 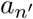, as well as a second-order neighbor of patient *a_n_*. Person *a_m_* is a first-order neighbor of patient 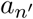, as well as a fourth-order neighbor of patient *a_n_*. As a result, it is more likely that *a_j_* is infected than *a_m_*; i.e., *p_j_* > *p_m_*.

## Discussion

Today, health systems around the world are prioritizing the use of tests for people who have either had direct contact with an infected person or are symptomatic. Some countries, such as South Korea, use extensive resources to test people randomly. On the other hand, there is a clear understanding that contact tracing technology, either by localization, or by contact monitoring using short range wireless communication, is a critical tool in dealing with rapidly evolving epidemics.

Our results show that the proper use of contact tracing must be complemented with a proper probabilistic analysis of the contact data (number of contacts, duration, etc.) to guide new tests of infection. The larger the testing capability, the more elaborate the algorithms must be to exploit the additional testing capacity.

We demonstrated that testing people using the methodology of a controlled experimental design in the spatio-temporal dimensions based on belief approximation exploits the available testing capacity much more efficiently. This results in a significant reduction of the burden on the healthcare system, the morbidity, and the negative economic and societal impact. The cost of expanding the number of tests is negligible compared to the tremendous positive advantages, and we encourage governments to invest resources in this effort. As shown for an advanced stage epidemic with 0.1% of the population already infected, the quarantine of large portions of the population (e.g., > 2%) can be reduced by over two months compared to random testing and by almost two weeks compared to simple neighbor testing, when up to 0.3% of the population is being tested every day. Similarly, the peak load on the health system can be reduced by 18% compared to testing randomly and by 10% compared to contact tracing with no controlled testing. Since the total morbidity is also lower, on one hand and also correlated with the death rate, many lives can be saved as well.

Similarly, it was shown that for early stage spread, using controlled testing can prevent the huge deterioration of the situation before it gets out of hand.

While controlled testing ideas can be applied under standard epidemiological investigations, without technological contact tracing, the delay involved and the inability of scaling to the millions, it clearly supports the use of technological contact tracing.

Another interesting observation is that random testing benefits significantly from very large numbers of tests, but that even testing up to 5% of the population daily still yields inferior results on both the deterministic and controlled testing policies. This underscores the importance of contact tracing and accurate probabilistic modeling.

Finally, some comments are in order regarding other factors. We used a false negative rate of 10 – 20%, but results scale favorably even with a 30% false negative rate. Similarly, our study assumed a 85% quarantine success rate. It was only mildly degraded when the quarantine rate went down to 70%. This shows that the method is robust to real world problems.

## Data Availability

This paper uses a stochastic agent-based model
with synthetic generated data.

## Supplementary materials

### Development of second-order approximation of the belief

The development of the second-order approximation of the belief used in ATI algorithm is described next. Let

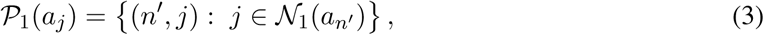

and

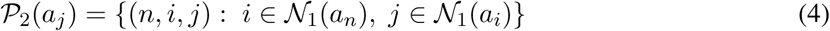

be the sets of all one-hop, and two-hop infection paths of person *j*, respectively. Then, the probability that person *a_j_* is infected through infection paths 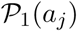 or 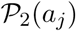 only is given by:

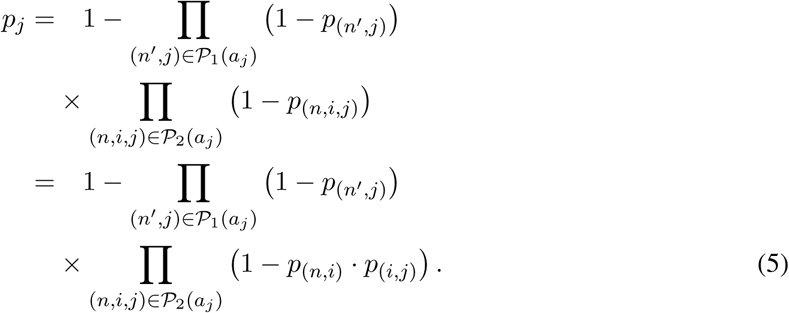

By assuming that *p_n_,_i_ <<* 1 for all *n, i*, we get:

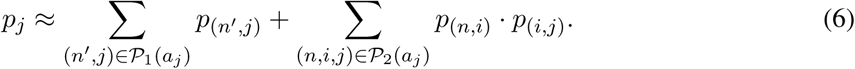

We use this approximation to update the beliefs over the population graph in the simulated models described next.

### Using simulated models to validate policy

#### Description of the graph topology

We simulated a COVID-19 outbreak environment with *N* people. The outbreak is initiated by *N_I_* infected people. In the simulations we set *N* = 100,000, and *N_I_* = 50, or *N* = 1, 000,000, and *N_I_* = 1,000. The degree distribution of the population graph captures different volume levels to model different types of social connections in the graph. For example, an office worker might have 25 close contacts, while a public service worker might have 200 close contacts. An example of the tail probability of the degrees in the population graph that we used in the simulations is illustrated in Fig. 5.

**Figure 5:**
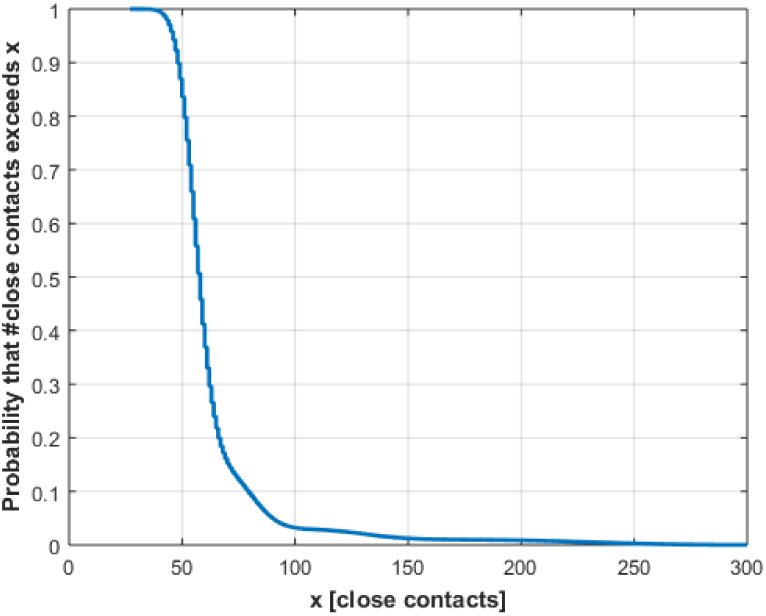
The probability that a person has greater than *x* close contacts (i.e., first-order neighbors) in the population graph.

#### Simulating symptomatic and asymptomatic people

An infected person can be asymptomatic or symptomatic. In the simulations we set 30% of people as asymptomatic. For asymptomatic people, symptoms do not appear and the only way to verify their condition is by testing them. For symptomatic people, we model the appearance time of symptoms by a Rayleigh distributed random variable with parameter 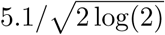, as illustrated in Fig. 6. This distribution models the appearance of symptoms mostly between day 2 and 11 from the day the person was infected, with a median of day 5.1, and a probability that symptoms will appear after day 11 of less 0.03. The distribution model and its parameters were chosen to fit the empirical incubation time recently reported in (23).

**Figure 6:**
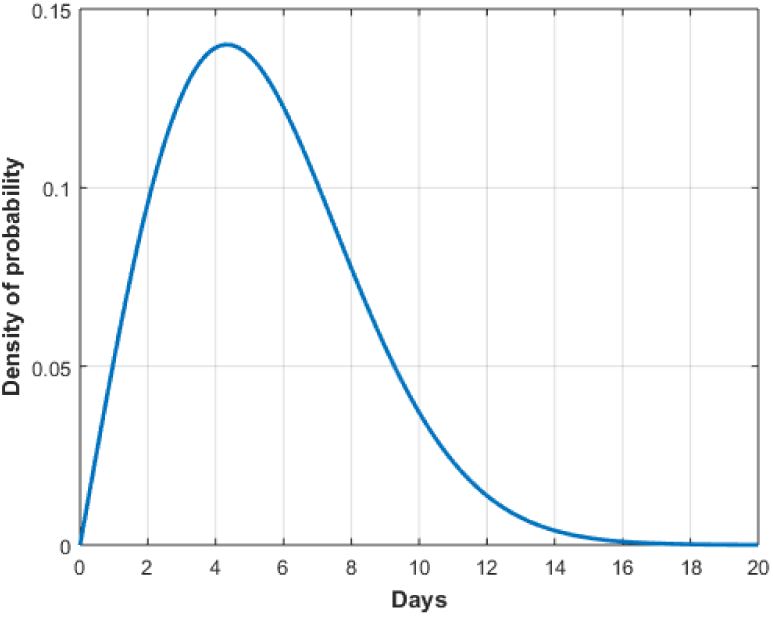
An illustration of the probability density function used to model the random time between the day a person was infected until symptoms appear (i.e., incubation time) in the simulation environment. The distribution is modeled by a Rayleigh distributed with parameter 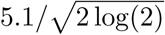, to fit the empirical incubation time recently reported in (23).

#### Transiting to static states

Static states in the simulations represent a state of a person who cannot infect other people or be infected by other people. This state captures a person who was infected and recovered, or sadly a person who was infected and died. The time between the day of infection and transitioning to the static state was set to 21 in the simulations.

#### Placing people in quarantine

Once a positive observation *o*(*a_n_*) for person *a_n_* is obtained, *a_n_* enters quarantine. This is done if symptoms appear for person *a_n_*, or if the person was scheduled for testing and a positive test result was obtained. The person enters quarantine until he or she transitions to the static state (recovers or dead), or after the recovery time has elapsed (set to 21 days in simulations) in the case of a false positive (we set 0.01 false positive rate in simulations). In addition, all its first-order neighbors 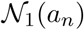 enter quarantine for at least *D* days, where *D* = 14 in the simulations, which is the typical isolation time used in many countries. People in quarantine cannot infect other people or be infected by other poeople. After 14 days, individuals in 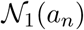 are released if symptoms have not appeared. To further model realistic situations, we assumed that *δ_s_* days elapse between the day symptoms appear and the day a person and his or her first-order neighbors enter quarantine. We set *δ_s_* = 3 in simulations. We also assumed that *δ_T_* days elapse between the day the test is performed and the result is received, and consequently the person and his or her first-order neighbors enter quarantine if positive. We set *δ_T_* = 1 in simulations.

#### False negatives in test results

Efficient COVID-19 tests should have low false negative rates to avoid releasing infected people and infecting other people. Today, COVID-19 tests suffer from roughly 0.1 – 0.2 false negative rates. To model this effect, we incorporated a parameter *FN* in the simulation environment. In the simulations, we set *FN* to 0.2.

#### Stochastic infections over the population graph

We set the infection probability oevr the graph by:

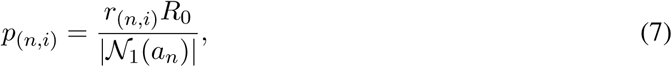

where 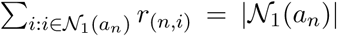, where the reproduction rate was set to *R*_0_ = 2.3 fixed for all the individuals in the simulations. This models the case where all people infect the same number of people *R*_0_ on average. It models the situation where people with a high degree of infection interacts with each person for less time, or use protective measures that reduce the infection risk (e.g., public service workers). Nevertheless, the simulator is generic and can be updated to simulate different infection probabilities for different people. The constants *r*_(*n i*)_ model the situation where the infection probability is not uniform among first-order neighbors. For example, being within 2 meters of a COVID-19 patient for a longer period of time increases the infection probability. In the simulations, we set *r*_(_*_n i_*_)_ to 0.25 for half of the first-order neighbors, and 1.75 for the other half, for each person.

#### Updating the belief in ATI algorithm

We simulated the suggested ATI algorithm using first-order and second-order approximations of the beliefs in Eq. (5), respectively, i.e.,

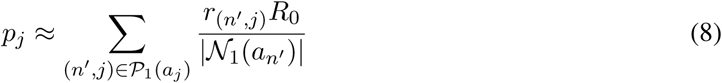

under the first-order ATI, and

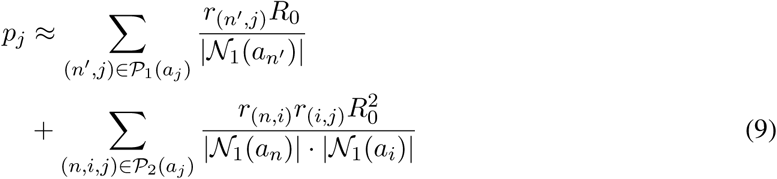

under the second-order ATI.

In addition, we used temporal updates of the belief. We reduced the belief of all people by multiplying it by a forgetting factor *α* at each iteration. This gives higher weight to recent observations. Specifically, at each iteration, we update the belief by:

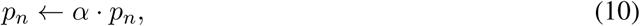

where ← denotes assignment notation. A forgetting factor of *α* = 0.75 demonstrated good performance. This fits a residual belief of approximatey 3% after 11 days, which fits the tail of the occurrence symptoms distribution.

Similarly, a negative test result should reduce the belief significantly. The proper way to do this is using a Bayesian update taking into account the false negative rate in the infected population, as well as the overall infection rate in the population. We found that this can be replaced quite well by a forgetting factor of *β* = 0.25 for a false negative rate of 0.1 – 0.2 without significant loss. Specifically, at each time a person *n* is tested and the result is negative, we update the belief by:

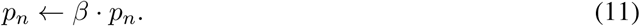

As usually done in adaptive algorithms, these parameters should be tuned, given the parameters of the environment.

#### Impact of quarantine efficiency on suppressing COVID-19

Next, we report the simulation results in addition to the results presented in the Performance Evaluation section. We simulated different values of quarantine efficiency, in which the quarantine was only adhered to 70%, 80%, 90%, 100% percent of the people required to be in quarantine, respectively. As a baseline to our suggested ATI method (first and second order), we simulated the NTI, RTI, and DTI methods, as described in the Performance Evaluation section. The results are presented in Tables 3, 4, and 5. Table 3 shows that the total infected people by NTI is between 1.6 and 9 times higher than ATI. Even using DTI results suffers from up to twice the morbidity as compared to ATI for a 70% quarantine success rate. It can be seen in Table 4 the peak number of infected people in NTI is up to 1.6 times higher than ATI. Even using DTI results suffers from up to 1.17 times the peak number of infected people as compared to ATI for a 70% quarantine success rate. Finally, in addition to the effectiveness of ATI in terms of reducing the burden on the health system, and reducing the total morbidity, Table 5 shows that ATI achieves these goals at a relatively small cost in terms of required qurantine days. Specifically, the total days lost for quarantine by NTI are between 1.6 and 8 times more than ATI, and even DTI requires between 1.17 and 2 times more quarantine days than ATI.

**Table 3:** Total # of infected people

| Quarantine success/method | NTI | RTI | DTI | 1st-order ATI | 2nd-order ATI |
| --- | --- | --- | --- | --- | --- |
| 70% | 8,256 | 7,516 | 1,962 | 909 | 911 |
| 80% | 2,042 | 1,658 | 620 | 445 | 432 |
| 90% | 756 | 612 | 385 | 318 | 334 |
| 100% | 434 | 423 | 314 | 266 | 274 |

**Table 4:** Peak # of infected people

| Quarantine success/method | NTI | RTI | DTI | 1st-order ATI | 2nd-order ATI |
| --- | --- | --- | --- | --- | --- |
| 70% | 701 | 648 | 507 | 431 | 442 |
| 80% | 509 | 488 | 408 | 349 | 344 |
| 90% | 407 | 374 | 331 | 293 | 308 |
| 100% | 340 | 335 | 295 | 261 | 270 |

**Table 5:** Total # of days in quarantine

| Quarantine success/method | NTI | RTI | DTI | 1st-order ATI | 2nd-order ATI |
| --- | --- | --- | --- | --- | --- |
| 70% | 4,118,872 | 3,816,708 | 1,046,777 | 509,401 | 508,265 |
| 80% | 1,298,703 | 1,055,431 | 395,188 | 286,326 | 278,950 |
| 90% | 549,878 | 446,522 | 279,297 | 230,953 | 242,417 |
| 100% | 352,256 | 342,529 | 255,518 | 217,295 | 223,400 |

## Footnotes

1 With more accurate contact tracing this graph becomes a weighted graph where the edges are the probability of mutual infection; e.g., parametrized by the duration of the contact

## References

1. Roy M Anderson, Christophe Fraser, Azra C Ghani, Christl A Donnelly, Steven Riley, Neil M Ferguson, Gabriel M Leung, Tai H Lam, and Anthony J Hedley. Epidemiology, transmission dynamics and control of SARS: The 2002-2003 epidemic. Philosophical Transactions of the Royal Society of London. Series B: Biological Sciences, 359(1447):1091–1105, 2004.

2. Roy M Anderson, Hans Heesterbeek, Don Klinkenberg, and T Déirdre Hollingsworth. How will country-based mitigation measures influence the course of the COVID-19 epidemic? The Lancet, 395(10228):931–934, 2020.

3. Herman Chernoff. Sequential design of experiments. The Annals of Mathematical Statistics, 30(3):755–770, 1959.

4. Kobi Cohen and Qing Zhao. Active hypothesis testing for anomaly detection. Information Theory, IEEE Transactions on, 61(3):1432–1450, 2015.

5. Kobi Cohen and Qing Zhao. Asymptotically optimal anomaly detection via sequential testing. Signal Processing, IEEE Transactions on, 63(11):2929–2941, 2015.

6. Duccio Fanelli and Francesco Piazza. Analysis and forecast of covid-19 spreading in china, italy and france. Chaos, Solitons & Fractals, 134:109761, 2020.

7. Luca Ferretti, Chris Wymant, Michelle Kendall, Lele Zhao, Anel Nurtay, Lucie Abeler-Dörner, Michael Parker, David Bonsall, and Christophe Fraser. Quantifying SARS-CoV-2 transmission suggests epidemic control with digital contact tracing. Science, 2020.

8. Christophe Fraser, Steven Riley, Roy M Anderson, and Neil M Ferguson. Factors that make an infectious disease outbreak controllable. Proceedings of the National Academy of Sciences, 101(16):6146–6151, 2004.

9. Giuseppe Gaeta. Asymptomatic infectives and *r*_0 for covid. *arXiv preprint arXiv:2003.14098*, 2020.

10. Laurent Hébert-Dufresne, Benjamin M Althouse, Samuel V Scarpino, and Antoine Allard. Beyond *r*_0: the importance of contact tracing when predicting epidemics. *arXiv preprint arXiv:2002.04004*, 2020.

11. Joel Hellewell, Sam Abbott, Amy Gimma, Nikos I Bosse, Christopher I Jarvis, Timothy W Russell, James D Munday, Adam J Kucharski, W John Edmunds, Fiona Sun, et al. Feasibility of controlling COVID-19 outbreaks by isolation of cases and contacts. The Lancet Global Health, 2020.

12. Bar Hemo, Tomer Gafni, Kobi Cohen, and Qing Zhao. Searching for anomalies over composite hypotheses. IEEE Transactions on Signal Processing, 68:1181–1196, 2020.

13. Herbert W Hethcote. The mathematics of infectious diseases. SIAM review, 42(4):599–653, 2000.

14. Javad Heydari and Ali Tajer. Controlled sensing for multi-hypothesis testing with co-dependent actions. In 2018 IEEE International Symposium on Information Theory (ISIT), pages 2321–2325. IEEE, 2018.

15. Javad Heydari, Ali Tajer, and H. Vincent Poor. Quickest linear search over correlated sequences. IEEE Transactions on Information Theory, 62(10):5786–5808, 2016.

16. T Déirdre Hollingsworth, Don Klinkenberg, Hans Heesterbeek, and Roy M Anderson. Mitigation strategies for pandemic Influenza A: Balancing conflicting policy objectives. PLoS computational biology, 7(2), 2011.

17. Boshuang Huang, Kobi Cohen, and Qing Zhao. Active anomaly detection in heterogeneous processes. IEEE Transactions on Information Theory, 65(4):2284–2301, 2019.

18. Alihan Hüyük and Cem Tekin. Analysis of thompson sampling for combinatorial multi-armed bandit with probabilistically triggered arms. *arXiv preprint arXiv:1809.02707*, 2018.

19. Omer Karin, Yinon M Bar-On, Tomer Milo, Itay Katzir, Avi Mayo, Yael Korem, Boaz Dudovich, Eran Yashiv, Amos J Zehavi, Nadav Davidovich, et al. Adaptive cyclic exit strategies from lockdown to suppress COVID-19 and allow economic activity. *medRxiv*, 2020.

20. Dhruva Kartik, Ashutosh Nayyar, and Urbashi Mitra. Fixed-horizon active hypothesis testing. *arXiv preprint arXiv:1911.06912*, 2019.

21. Fuminori Kato, Kei-ichi Tainaka, Shogo Sone, Satoru Morita, Hiroyuki Iida, and Jin Yoshimura. Combined effects of prevention and quarantine on a breakout in sir model. Scientific reports, 1:10, 2011.

22. Adam J Kucharski, Timothy W Russell, Charlie Diamond, Yang Liu, John Edmunds, Sebastian Funk, Rosalind M Eggo, Fiona Sun, Mark Jit, James D Munday, et al. Early dynamics of transmission and control of COVID-19: A mathematical modelling study. The lancet infectious diseases, 2020.

23. Stephen A Lauer, Kyra H Grantz, Qifang Bi, Forrest K Jones, Qulu Zheng, Hannah R Meredith, Andrew S Azman, Nicholas G Reich, and Justin Lessler. The incubation period of coronavirus disease 2019 (covid-19) from publicly reported confirmed cases: estimation and application. Annals of internal medicine, 2020.

24. Dror Meidan, Reuven Cohen, Simcha Haber, and Baruch Barzel. An alternating lock-down strategy for sustainable mitigation of COVID-19. *arXiv preprint arXiv:2004.01453*, 2020.

25. Mohammad Naghshvar and Tara Javidi. Active sequential hypothesis testing. The Annals of Statistics, 41(6):2703–2738, 2013.

26. Yixiang Ng, Zongbin Li, Yi Xian Chua, Wei Liang Chaw, Zheng Zhao, Benjamin Er, Rachael Pung, Calvin J Chiew, David C Lye, Derrick Heng, et al. Evaluation of the effectiveness of surveillance and containment measures for the first 100 patients with COVID-19 in Singapore–January 2–February 29, 2020. 2020.

27. Sirin Nitinawarat, George K Atia, and Venugopal V Veeravalli. Controlled sensing for multihypothesis testing. IEEE Transactions on Automatic Control, 58(10):2451–2464, 2013.

28. Sirin Nitinawarat and Venupogal V Veeravalli. Controlled sensing for sequential multihypothesis testing with controlled Markovian observations and non-uniform control cost. Sequential Analysis, 34(1):1–24, 2015.

29. Rahul Potluri and Deepthi Lavu. Making sense of the global coronavirus data: The role of testing rates in understanding the pandemic and our exit strategy. *Available at SSRN 3570304*, 2020.

30. Herbert Robbins. Some aspects of the sequential design of experiments. Bulletin of the American Mathematical Society, 58(5):527–535, 1952.

31. Alfonso J Rodriguez-Morales, K Bonilla-Aldana, R Ruchi Tiwari, Ranjit Sah, Ali A Rabaan, and Kuldeep Dhama. COVID-19, an Emerging Coronavirus Infection: Current Scenario and Recent Developments-An Overview. Journal of Pure and Applied Microbiology, 14:6150, 2020.

32. Marcel Salathé, Christian L Althaus, Richard Neher, Silvia Stringhini, Emma Hodcroft, Jacques Fellay, Marcel Zwahlen, Gabriela Senti, Manuel Battegay, Annelies Wilder-Smith, et al. COVID-19 epidemic in Switzerland: On the importance of testing, contact tracing and isolation. Swiss medical weekly, 150(1112), 2020.

33. Shai Shalev-Shwartz and Amnon Shashua. Can we contain COVID-19 without locking-down the economy? Technical report, Center for Brains, Minds and Machines (CBMM), 2020.

34. Aristomenis Tsopelakos, Georgios Fellouris, and Venugopal V Veeravalli. Sequential anomaly detection with observation control. In 2019 IEEE International Symposium on Information Theory (ISIT), pages 2389–2393. IEEE, 2019.

35. Hoi-To Wai, Anna Scaglione, Baruch Barzel, and Amir Leshem. Joint network topology and dynamics recovery from perturbed stationary points. IEEE Transactions on Signal Processing, 67(17):4582–4596, 2019.

36. Hoi-To Wai, Anna Scaglione, and Amir Leshem. Active sensing of social networks. IEEE Transactions on Signal and Information Processing over Networks, 2(3):406–419, 2016.

37. World Health Organization. Considerations for quarantine of individuals in the context of containment for coronavirus disease (COVID-19): interim guidance, 19 March 2020. Technical report, World Health Organization, 2020.

38. Qing Zhao. Multi-armed bandits: Theory and applications to online learning in networks. Synthesis Lectures on Communication Networks, 12(1):1–165, 2019.

